# Outcomes of a multi-objective diet optimization model as a basis for food-based dietary guidelines

**DOI:** 10.64898/2026.11.19.26368467

**Authors:** Anne C. Schäfer, Heiner Boeing, Johanna Conrad, Rozenn Gazan, Kurt Gedrich, Hans Hauner, Anja Kroke, Jakob Linseisen, Stefan Lorkowski, Lukas Schwingshackl, Florent Vieux, Kiran Virmani, Ute Nöthlings, Bernhard Watzl

## Abstract

**Background:** While nutrient adequacy has been at the core of food-based dietary guidelines (FBDG) for Germany since their beginning, aspects of health-promoting and more sustainable diets have not yet been integrated into their quantitative derivation.

**Objective:** To consider multiple aspects relevant to FBDGs simultaneously via a multi-objective diet optimization model and to conceptualize an approach to transfer the outcomes into a basis for FBDGs, i.e. food guide and messages, on the example of adults in Germany.

**Methods:** The optimization model included FoodEx2 food codes as decision variables (n = 593, level 4), constraints on nutrient intake recommendations, acceptability, and agronomic dependencies, and a multi-objective function that simultaneously minimized diet-related disease burden (disability-adjusted life years, DALYs), environmental impact (greenhouse gas emissions (GHGE) and land use), and deviation from the observed diet. Ten model variants were evaluated using five different weighting schemes for the components of the objective function and two scenarios. For the transfer, optimization results were first further aggregated, then transferred into circular charts for the food guide and into servings for the FBDG messages.

**Results:** Meeting all constraints, the optimized diets reduced DALYs by 39-77 %, GHGE by 40-48 % and land use by 44-57 %. Compared with the observed diet, optimized diets were characterized by a higher share of plant-based foods in the total quantity of foods (66-70 % vs. 55 %), energy contribution (77-79 % vs. 58 %), and protein contribution (63-68 % vs. 40 %). The optimized food group quantities (coefficient of variation ranging from 0 to 1.0), the circular charts, and the servings showed high similarity across the ten model variants.

**Conclusions:** Our findings demonstrate that diet optimization outcomes can serve as a basis for health-promoting and environmentally more sustainable FBDGs. The model’s flexibility allows future improvements in data quality and availability to be incorporated.

## Introduction

Diet-related noncommunicable diseases such as obesity, type 2 diabetes, and cardiometabolic diseases constitute major public health concerns worldwide [1]. Another pressing issue is that especially Western dietary patterns are a major driver of various environmental challenges [2]. Food-based dietary guidelines (FBDGs) advise on healthy diets and should be tailored to country-specific health problems [3]. They further consider environmental and food system aspects while ensuring compatibility with the habitual national diet [4–6]. Many countries provide both a food guide, which visually represents the recommended proportions of different food groups, and quantitative and qualitative messages within their FBDGs to communicate which food groups should be prioritized and in what amounts [4,7].

Deriving FBDGs that integrate multiple aspects simultaneously may benefit from approaches such as diet optimization which can address these complexities [8–10]. Basically, diet optimization models consist of decision variables (foods or food groups), constraints (lower and upper bounds and target values that define the feasible solution space), and an objective function that identifies optimal diets within that defined space. This method has been successfully applied in the derivation of various national FBDGs [10–18]. To use the outcomes of diet optimization models - optimized quantities for up to several hundred of food groups - as a basis for FBDGs, they must be transferred into simpler formats for public communication. This includes the food guide, as well as quantitative and qualitative messages. However, although some examples of transferring optimization results into food guides exist [15–17], detailed information on the transfer process, especially for the messages, is rare.

In Germany, the FBDG 2013 by the German Nutrition Society (DGE) [19,20] offered health benefits [21,22], but the environmental impact would have exceeded planetary boundaries if adopted by the global population [23]. Consequently, a new edition of the FBDG for Germany should integrate both aspects of health and environmental sustainability [24]. Similarly, the DGE aimed to base its FBDG on the outcomes of an optimization model and establish a consistent approach for the transfer of these outcomes into their FBDG. Investigations into methodological considerations such as the choice of nutrient goals, decision variables, the mathematical form of the objective function [25], and weighting schemes (WS) of the multi-objective function [26] provided the foundation of the optimization model used in this study.

This study aimed to develop diets that simultaneously consider aspects of health, environmental sustainability, and acceptability via a multi-objective diet optimization model and to transfer the resulting optimized diets into quantitative and qualitative recommendations, using the FBDG for adults in Germany as an example.

## Methods

### Decision variables

**Figure 1** provides an overview of the diet optimization model (version 24.0.0). To define the food groups used as decision variables, we used the hierarchical food group classification system FoodEx2 by the European Food Safety Authority (EFSA) [27], which was adjusted to differentiate between refined and whole grain and between processed meat and unprocessed meat [25]. FoodEx2 consists of seven levels, with Level 1 being the most aggregated. Since the introduction of an agronomic dependency on red meat (see below) required a more detailed classification, Level 4 was selected for the decision variables [25]. The optimization model consisted of 593 FoodEx2 food groups (decision variables), including discretionary food groups.

**Figure 1:**
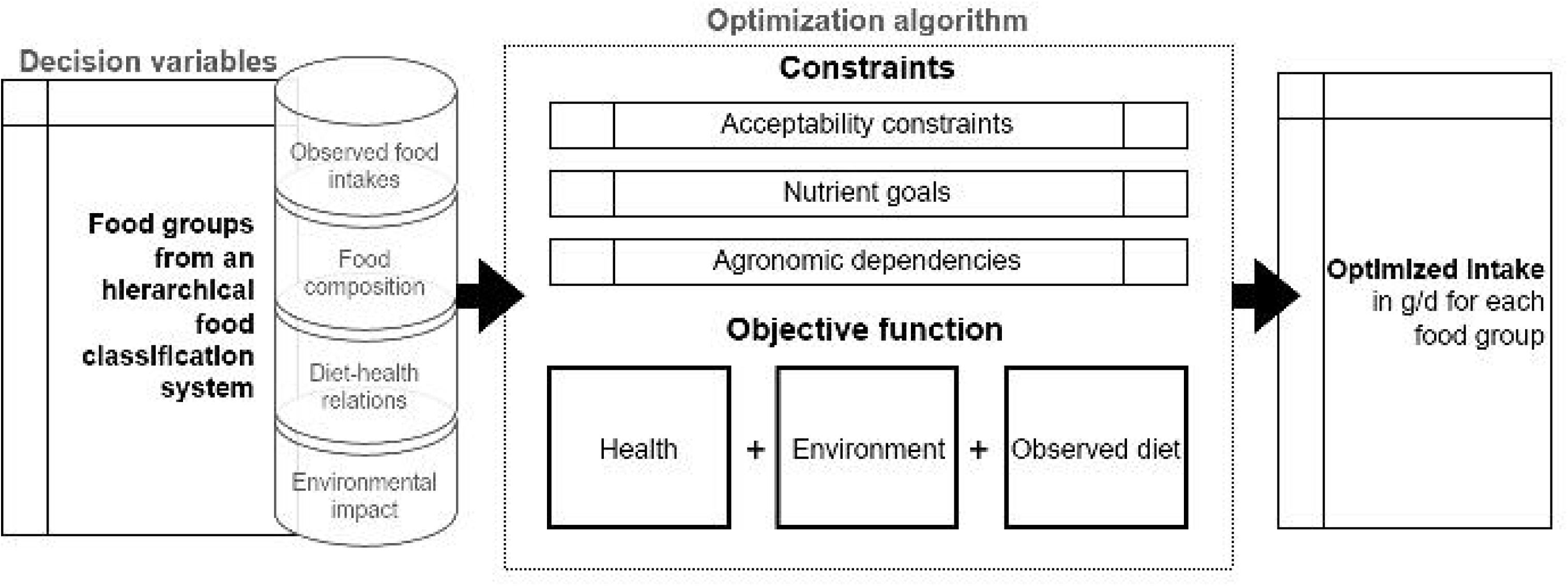
Overview of the diet optimization model (version 24.0.0) for deriving the FBDG for Germany.

Different datasets accounting for health, environmental impact, and aspects of practical relevance (observed diet and agronomic dependencies) were assigned to the decision variables. The observed diet data were based on representative food intake data for German adults aged 18-65 years, from the most recent German National Nutrition Survey II (NVS II) (2005-2007) [28–30]. We retrieved the data from the EFSA Comprehensive European Food Consumption Database [31,32]. For food composition data, we used the German food composition database (Bundeslebensmittelschlüssel version 3.02) [33], and for free sugars, we used data from LEBTAB [34]. Disability-adjusted life years (DALYs) from both the Global Burden of Disease Study [1,35] and Schwingshackl et al. [36] were used as indicators for diet-health relations. Following a previously described methodology [26], diet-related DALYs for each food group’s food intake level were estimated by linear interpolation between the observed food intake, associated with the reported diet-related DALYs, and the optimal food intake (theoretical minimum risk exposure level), associated with zero DALYs.

Environmental impact data on greenhouse gas emissions (GHGE in kg CO_2_ equivalents) and land use (m^2^*year) were obtained from the SHARP-Indicators Database, which contains life cycle assessment data including production, packaging, transport, and cooking [37,38].

### Optimization algorithm

The solution space was framed by constraints on acceptable food group quantities (acceptability constraints) at the 5^th^ and 95^th^ percentiles of observed food intake [25] (Figure 1). Additional constraints on food group quantities were set for liver (mean observed intake across all individuals as the upper bound) and for the decision variables of composite dishes and sauces and condiments (mean observed intake across all individuals as the upper bound, scaled towards the energy goal of the optimized diet) [25]. Adequate nutrient supply was ensured by setting nutrient goals based on the mean of the dietary reference values for the different age and sex groups among adults aged 18–65 years, weighted according to their representation in the population [25]. Further constraints were set on agronomic dependencies. They considered the co-production of foods and were introduced for the following food combinations: a ratio of 100:2 for milk to beef production, which corresponds to a circular economy on grassland [39]; a ratio between 100:80 and 100:120 for unprocessed red meat to processed meat [40]; between 100:2.5 and 100:5 for milk to butter [41]. To calculate milk equivalents from milk and dairy products, we used conversion factors of 1 for milk, 7.2 for cheese, and 1.4 for any other milk product (e.g., cream) [42].

The multi-objective function consisted of three components, including “health,” i.e., the minimization of disease burden (DALYs from Schwingshackl et al. [36] and Global Burden of Disease [1,35], [26]), “environment,” i.e., the minimization of environmental impact (GHGE and land use) [37], and “observed diet,” i.e., the minimization of the relative deviation from the mean observed food intakes across all individuals from the NVS II [28–30] (Figure 1). The multi-objective function was defined as a weighted-sum function in which the three components were standardized to ensure comparability [26].

We analyzed ten model variants applying five WS in two scenarios as the scientific basis for the German FBDG (**Table 1**). In each model, we assigned the observed diet a weight of 40 %, which was empirically determined to be low enough to enable a 45 % (−50 %, -40 %) reduction in GHGE [26,43]. The health and environment components were each assigned a weight of 30 % (Table 1, WS 3) as a starting point. We further introduced four WS involving 5 % steps in the weighting of health and environment (WS 1, 2, 4, and 5). WS 1-5 were applied in the first scenario without additional constraints and in the second scenario with a constraint corresponding to 300 g of meat per week, including red meat, processed meat, and poultry (FoodEx2 code: A01QR). The second scenario followed the German FBDG 2013 recommendation to eat no more than 300 g of meat, including processed meat, per week [19]. Modelling was performed using R version 4.1.3 [44] and the R Optimization Infrastructure package version 0.3-3 [45]. The linear programming problem was solved using the lpSolve R package, which uses the simplex algorithm [46]. The data was managed with MySQL Workbench version 8.0.

**Table 1:**
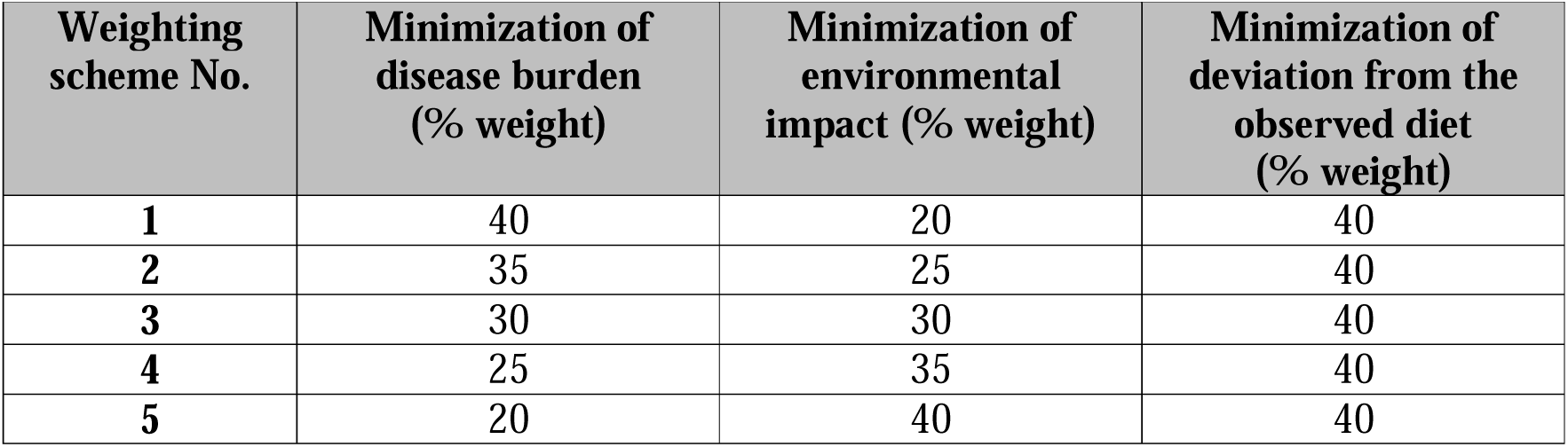
Weighting schemes for the multi-objective function of the diet optimization model.

| **Weighting scheme No.** | **Minimization of disease burden (% weight)** | **Minimization of environmental impact (% weight)** | **Minimization of deviation from the observed diet (% weight)** |
|---|---|---|---|
| **1** | 40 | 20 | 40 |
| **2** | 35 | 25 | 40 |
| **3** | 30 | 30 | 40 |
| **4** | 25 | 35 | 40 |
| **5** | 20 | 40 | 40 |

### Outcome measures

For ease of presentation and communication of the optimization results, the 593 decision variables were aggregated a posteriori into 18 food groups and an energy share for discretionary food groups [25]. To compare the observed and optimized diets, we described the respective energy and nutrient contents as well as the contributing food groups. The degree of similarity in food quantity was examined by calculating the coefficient of variation across the ten model variants. To assess the health implications of the optimization results, we calculated the relative reduction in DALYs for the optimized diets compared with the observed diet [26] and checked for the fulfillment of nutrient constraints. Regarding environmental impact, we compared the GHGE and land use of the optimized diets with those of the observed diet. Regarding the acceptability and practicality of the optimized diets, we checked whether the constraints on acceptability as well as agronomic dependencies were fulfilled.

### Transfer

**Figure 2** depicts the steps taken to transfer the outcomes of the diet optimization model into the food guide, which visualizes the proportions of the different food groups, and into quantitative and qualitative messages. First, for both the food guide and the messages, food groups were further aggregated beyond the initial aggregation into the 18 food groups.

**Figure 2:**
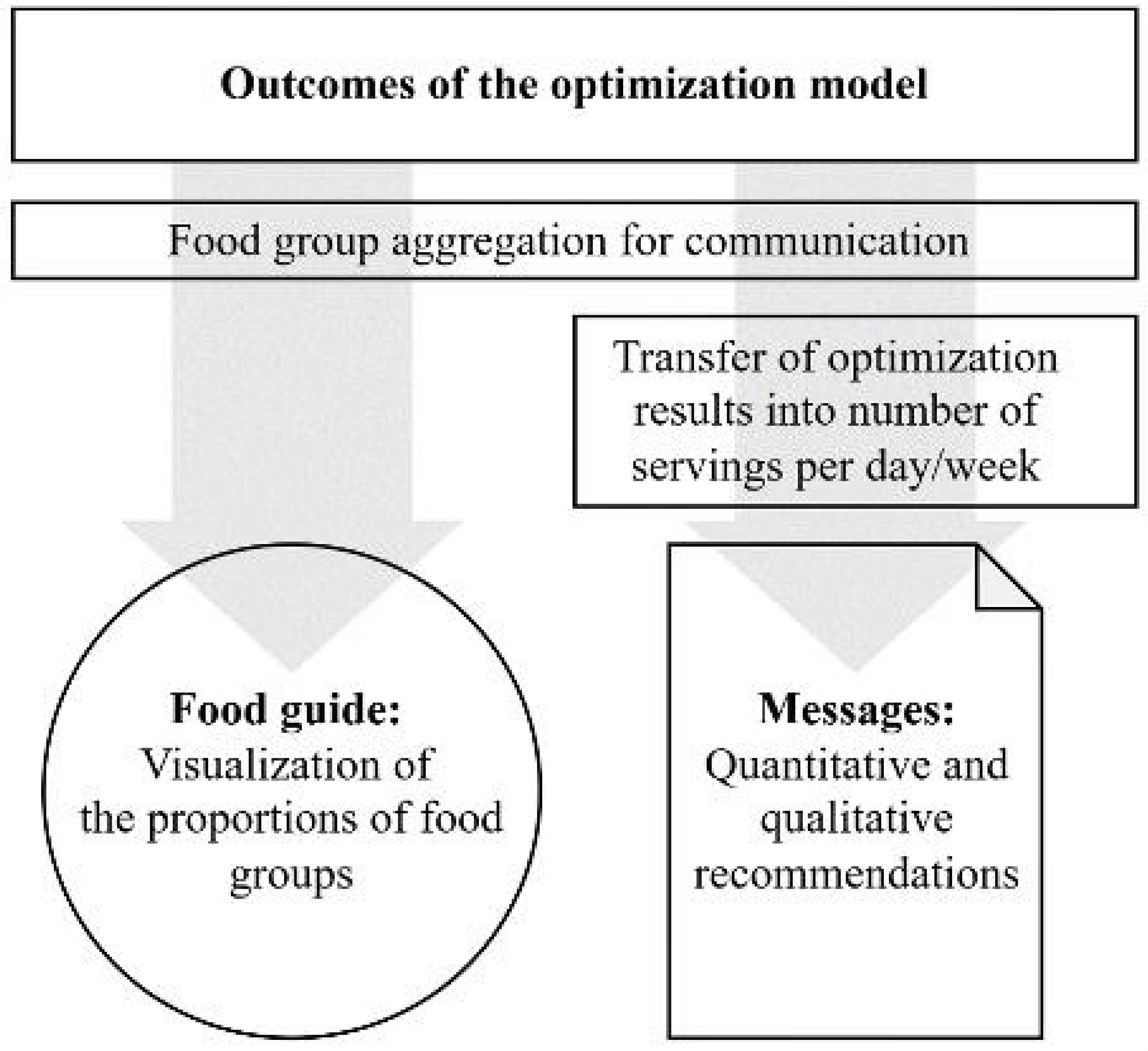
Process for the transfer of optimization results into food guide and messages.

To determine the proportions displayed in the food guide, the optimized food quantities from each model were converted separately into graphical representations. The similarity of the graphical representations was assessed both visually, by comparing the overlap of food group segments, and quantitatively, by evaluating standard deviations across different combinations of model variants.

For the transfer into FBDG messages, the daily or weekly number of servings for each food group was calculated as follows: After the aggregation of the food groups, the optimized food quantities of the ten model variants were divided by the predefined portion sizes (see Results, Table 4). The results were rounded to the nearest integer, with values of 0.5 or greater rounded up. The consumption interval was daily for all food groups, except for fruit and vegetable juices, legumes, potatoes, eggs, fish, meat, and processed meat, for which a weekly interval was more appropriate. For legumes, the fresh weight was used to derive servings and messages (conversion factor from dry weight to fresh weight = 1.8). The servings were then used to derive parts of the quantitative and qualitative recommendations within the messages that described the developed dietary pattern.

For more details on the general process to derive the DGE’s FBDG 2024, see **Supplementary Methods**.

## Results

### Health and environmental implications of the observed diet in Germany

The observed diet (NVS II) showed a mean energy intake of 2,277 kcal/d with 45 En% (percent of total energy intake) from carbohydrates, 38 En% from fat, and 14 En% from protein (including discretionary foods). Grains and grain-based products (29 %) and milk and dairy products (10 %) were the major contributors to energy intake. The observed diet exceeded, among other recommendations, the recommended amounts of salt, free sugars, and saturated fatty acids, whereas the dietary reference values for some recommendations, e.g. dietary fiber, were not achieved. See **Supplementary Table 1** for more details on energy and nutrient contents.

The disease burden related to the observed diet for adults in Germany was 2,073,195 DALYs according to the Global Burden of Disease Collaborative Network [35] and 2,192,961 DALYs according to Schwingshackl et al. [36]. The environmental impact was 6.18 kg CO_2_ equivalents/d and 7.48 m^2^*year/d of land use (Supplementary Table 1). Milk and dairy products (13 %), red meat (13 %), composite dishes (12 %), processed meat (10 %), and refined grains (10 %) accounted for approximately 60% of GHGE. Red meat (18 %), processed meat (13 %), composite dishes (16 %), and refined grains (15 %) accounted for approximately 60 % of land use (**Supplementary Table 2**).

### Optimized diets

The optimized food intakes of the ten model variants (five WS and two scenarios) are presented in **Table 2**. All constraints (acceptability, nutrient goals, and agronomic dependencies) were fulfilled for each optimized diet. The energy content was 2,029 kcal/d with 55-56 En% from carbohydrates, 29 En% from fat, and 14 En% from protein across the ten model variants. Details of the energy and macro- and micronutrient contents are shown in Supplementary Table 1.

**Table 2:**
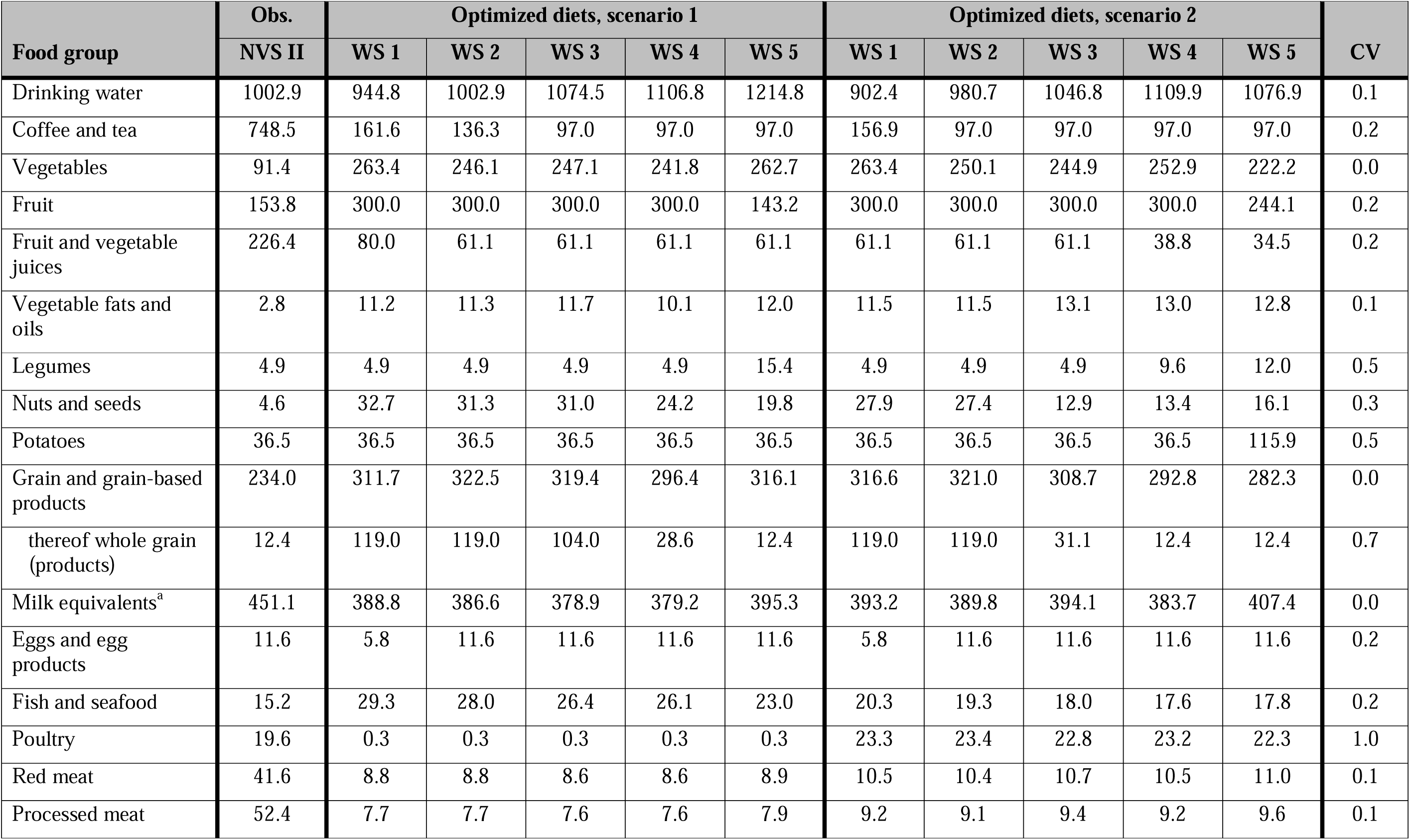
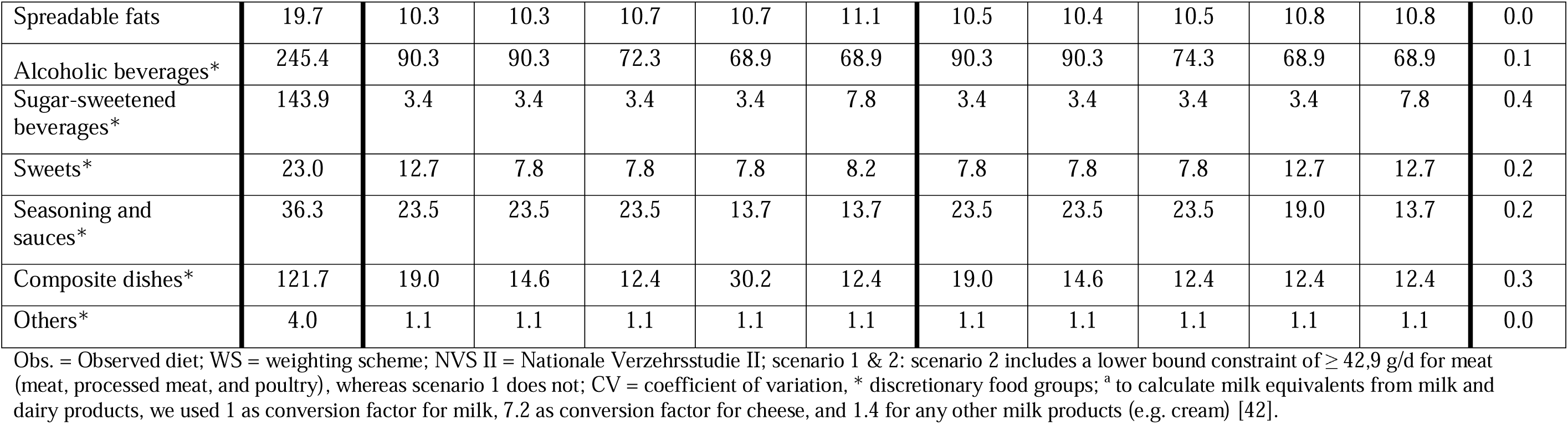
Observed and optimized food intakes for the 18 food groups and for 6 discretionary food groups (g/d).

The optimized diets were characterized by higher quantities of vegetables, fruit (except in WS 5, scenario 1), whole grains, fish, and vegetable oils, and lower quantities of red meat and processed meat than the observed diet. The quantities of potatoes, egg, and milk equivalents remained approximately the same (Table 2). Discretionary foods decreased from 22 En% in the observed diet to 7-9 En% in the optimized diets. The changes in food quantities across the ten optimized diets were also reflected in the proportion of plant-based foods in the total quantity of foods (66-70 % vs. 55 % in the observed diet), their contribution to total energy (77-79 % vs. 58 % in the observed diet), and their contribution to total protein (63-68 % vs. 40 % in the observed diet) (**Supplementary Table 3**).

The optimized food group quantities (e.g., vegetables, fruits, potatoes, eggs, red meat, and processed meat) were generally stable across the five WS in both scenarios, as shown by the low coefficients of variation (CV) (Table 2). Exceptions were found for poultry, which had an optimized quantity of approximately 23 g/d in scenario 2, compared with 0 g/d in scenario 1 (CV = 1.0). The introduction of a lower bound on meat in scenario 2 also led to approximately 10 g/d less fish than in the scenario 1 model variants. For whole grains, the optimized quantity varied between 12 g/d and 119 g/d (CV = 0.7) across the ten models. Finally, the model variants with WS 4 and WS 5 deviated from the other model variants in a few food groups. One example was legumes, for which a higher quantity of 10–15 g/d was used in the models with greater environmental weighting (WS 4 and WS 5), compared to 5 g/d in the other model variants (CV = 0.5).

The optimization model identified solutions that reduced diet-related DALYs by at least 39 % (WS 5, scenario 1) up to 77 % (WS 1, scenario 1) (**Table 3**). The DALY calculations are presented in **Supplementary Table 4**. Regarding the environmental impact, optimized diets achieved a 40 % (WS 1, scenario 2) to 48 % (WS 4 and WS 5, scenario 1) reduction in GHGE, remaining within the 40–50 % confidence interval defined by the Intergovernmental Panel on Climate Change [43]. Land use was reduced by 44 % (WS 1, scenario 2) to 57 % (WS 4 and WS 5, scenario 1). Similar food groups contributed to these reductions across all the model variants. Higher quantities of whole grains and lower quantities of processed meat, red meat, and sugar-sweetened beverages contributed most to the decrease in DALYs (**Supplementary Table 5**). Lower quantities of composite dishes, red meat, and processed meat contributed most to the reduction in the environmental impact (Supplementary Table 2).

**Table 3:** Reduction (%) of disability-adjusted life years, greenhouse gas emissions, and land use of the optimized diets compared to the observed diet.

|  | **Optimized diets, scenario 1** |  |  |  |  | **Optimized diets, scenario 2** |  |  |  |  |
|---|---|---|---|---|---|---|---|---|---|---|
| **Indicator** | **WS 1** | **WS 2** | **WS 3** | **WS 4** | **WS 5** | **WS 1** | **WS 2** | **WS 3** | **WS 4** | **WS 5** |
| DALYs (mean between both DALY sources) | -77 | -76 | -73 | -55 | -39 | -74 | -73 | -49 | -47 | -43 |
| GHGE (kg CO<sub>2</sub> equivalents/d) | -43 | -45 | -46 | -48 | -48 | -40 | -42 | -44 | -45 | -47 |
| Land use (m<sup>2</sup>*year/d) | -48 | -48 | -52 | -57 | -57 | -44 | -45 | -50 | -53 | -53 |

WS = weighting scheme; scenario 1 & 2: scenario 2 includes a lower bound constraint of ≥ 42,9 g/d for meat (meat, processed meat, and poultry), whereas scenario 1 does not; DALYs = disability-adjusted life years, GHGE = greenhouse gas emissions.

**Table 4:**
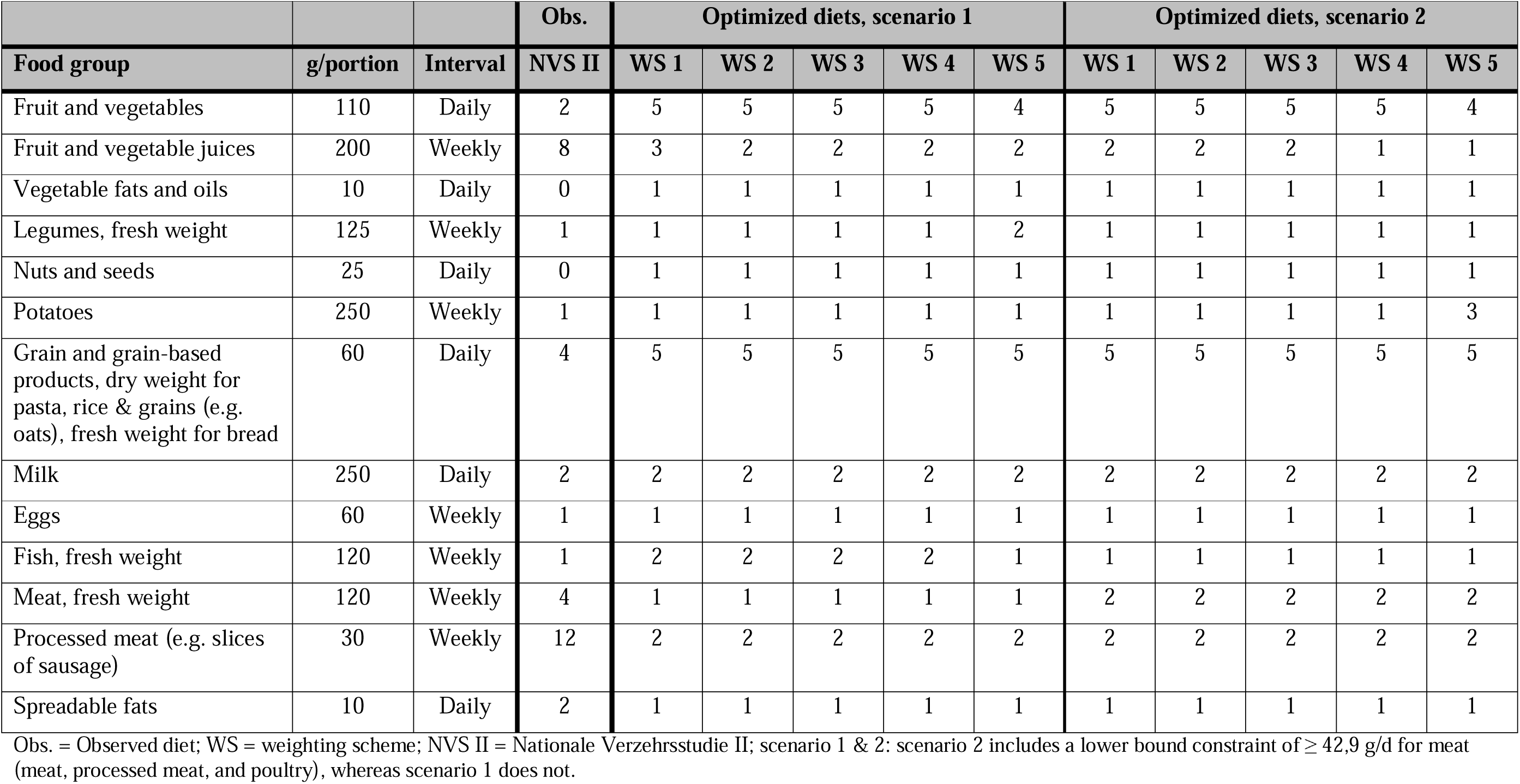
Predefined portion sizes of food groups and servings based on optimization results.

### Transfer of diet optimization outcomes into food guide and messages

For visualization as a food guide, the DGE decided to use a circle with pie chart segments, as this allows the relative proportions of food groups to be displayed. This type of food guide has been used in Germany since 1956. Following the food group aggregation, the DGE nutrition circle (**Figure 3)** was divided into seven segments: (1) fruit and vegetables, including juices; (2) legumes (dry weight) and nuts; (3) grains, grain-based products, and potatoes; (4) oils and fats; (5) milk and dairy products; (6) meat, fish, and eggs; and (7) beverages. For the food group milk and dairy, the optimized quantities of milk equivalents were converted into quantities of milk, milk products, and cheese, according to the proportions observed in NVS II. It should be noted that there are dried fruits and vegetables in segment (1) and that segment (3) consists of a mixture of bread, dried noodles/rice, and grains. The order of the food groups reflects the proportion of plant-based foods (segments 1-3) and animal-source foods (segments 5 and 6). Segment (4) is a mixture of plant-based and animal-sourced oils and fats.

**Figure 3:**
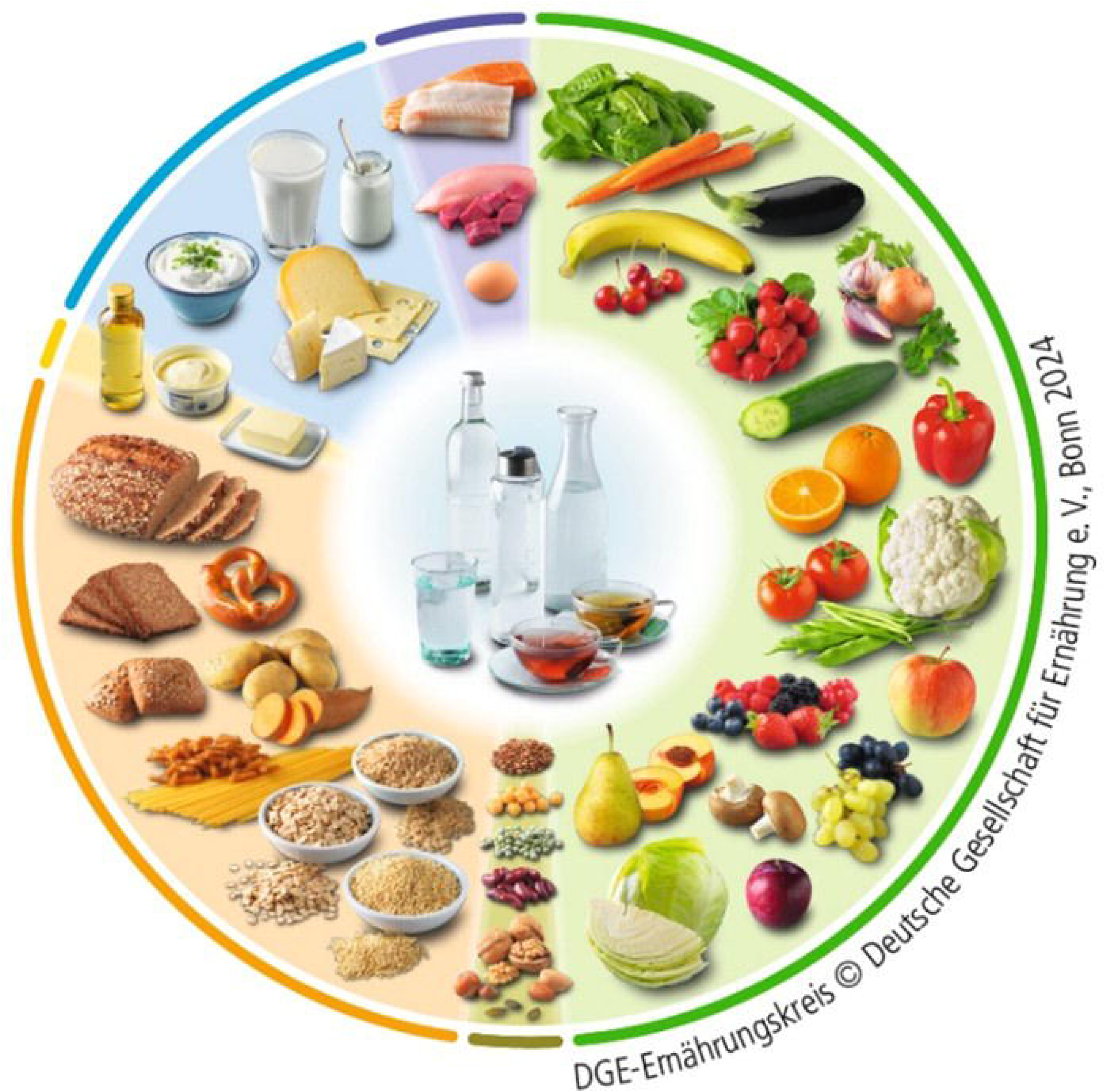
Diet optimization results transferred into a food guide: the DGE nutrition circle 2024.

The comparison of the ten model variants showed that WS 1 through 4 were very similar (SD ranging from 0.1 to 2.9), whereas the proportions in WS 5 differed due to the lower quantity of fruit and vegetables (> 100 g/d less; SD for WS 1 between 0.1 and 0.7; see Supplementary Table 6). Therefore, the circular charts of WS 1–4 in both scenarios were used as the basis for the DGE nutrition circle. The slight variation between these models is illustrated by a smooth transition between segments.

**Table 4** shows the predefined portion sizes and number of servings for 13 food groups derived from the model results. The number of servings was the same across most food groups and model variants: one serving of vegetable fats and oils per day, one serving of nuts and seeds per day, five servings of grains and grain-based products per day, two servings of milk and dairy per day, one egg per week, two servings of processed meat per week, and one serving of spreadable fats per day. For fruit and vegetables, fruit and vegetable juices, legumes, and potatoes, the results of WS 1 and 5 (and in the case of juices WS 4) partly differed from the other results by one serving more or less (in the case of potatoes, two servings more). Fish and meat were complementary, showing two servings of meat and one serving of fish per week in scenario 1, and vice versa in scenario 2. In scenario 1, WS 5 was an exception, with one serving of meat per week.

For the formulation of the messages, a “more is better” formulation was applied to fruit and vegetables, legumes, and whole grain, whereas a “less is better” formulation was applied to meat and discretionary foods. All model variants informed the derivation of recommendations within the messages. For example, the higher legume quantities in model variants 4 and 5, together with evidence of the beneficial role of legumes in sustainable diets, informed the recommendation to “eat *at least* one serving of legumes per week”.

## Discussion

In this study, we successfully developed diets that integrated multiple aspects of health and sustainability while remaining as close as possible to the observed diet, and transferred the optimization results into quantitative and qualitative recommendations in a consistent process. With the simultaneous consideration of diet-health relations, environmental aspects, and the observed diet, our optimization model showed robust results across the model variants and achieved substantial reductions in diet-related disease burden (DALYs reduced by 39-77 %) and environmental impact (GHGE reduced by 40-48 % and land use by approximately 44-57 %). In general, the optimized diets were characterized by a high proportion (more than three-quarters) of plant-based foods, such as fruit, vegetables, grains, and nuts and seeds, and lower quantities of processed meat, red meat, and sugar-sweetened beverages compared with the observed diet. The synergies of such plant-based diets for health and the environment, especially including the reduction in red meat, have been abundantly reported [47–50].

Our study demonstrated that most food groups showed little variation in food quantities for the ten model variants. The model stability, and respective robustness, was in line with our expectations, since the constraints and objective function of the model variants were similar. Furthermore, in previous analyses, we identified that the inclusion of multiple heterogeneous indicators and the assignment of a fixed weight to the observed diet are stabilizing factors [25,26]. Our results also align with previous studies of optimization models, which found that diet modelling is highly sensitive to approaches regarding the deviation from the observed diet [49] and that a combination of diverging components strengthens model stability [47].

However, some food groups were more sensitive to changes in the weighting of the health and environmental components in the objective function. For instance, while the quantity of grains was similar across the ten model variants, possibly reflecting the requirements for energy, fiber, and micronutrients, the proportion of whole grains within the grain group varied. In the model variants with a higher weighting on health, the optimized quantity was 119 g/d, corresponding to the theoretical minimum risk exposure level of whole grain [36]. In the model variants with lower weighting for health, the quantity of whole grains was lower; for example, reaching the observed intake level of 12 g/d in WS 5. These differences cannot be explained by nutritional requirements but rather by the nature of the multi-objective function, which seeks an optimum along a Pareto front [51–53]. Whole grains are characterized by a discrepancy between their high health value and their low observed intake in the population. This demonstrates that multi-objective optimization models can encounter difficulties in achieving consistent results when individual components of the objective function are in greater conflict with each other [52]. This highlights the need to carefully examine the input data and potentially combine the indicators into clusters to decrease the complexity of the optimization model [53].

The varying quantities of whole grain were also the main reason for the magnitude of variation in DALY reduction between the model variants (Δ = 39 % between WS1 (−77 %) and WS5 (−38 %), scenario 1). In another analysis (Supplementary Table 4a), we used the quantity of whole grains of 119 g/d from model variants 1 and 2 for all model variants to calculate DALYs. This resulted in less variation between models (Δ = 18 % between WS1 (−77 %) and WS5 (−59 %), scenario 1). Considering the strong effect of a diet low in whole grains on the diet-related burden of disease [1,36], an intake of 119 g whole grains/d was used as the basis for the German FBDG [54].

The introduction of the meat constraint (red and processed meat and poultry) in scenario 2 led to another variation in optimization outcomes. This resulted in an increase in the quantity for poultry, while fish decreased by one serving per week compared with scenario 1. These differences reflect the interplay of all model objectives and constraints: micronutrient requirements were already met by other food groups, whereas constraints such as saturated fatty acids, agronomic dependencies, and environmental impacts may have limited further increases in red and processed meat. This illustrates that the changes in individual food groups must be interpreted within the context of the entire diet. The findings regarding meat and fish are consistent with the Nordic Nutrition Recommendations 2023, which recommend keeping red meat and poultry intake low and replacing them with plant foods and sustainably sourced fish [55].

The outcomes of the multi-objective diet optimization model shown in the present study contributed directly to the DGE nutrition circle (food guide), illustrating the proportions of food groups and food quantities communicated through the messages. The DGE nutrition circle and the messages “Eat and drink well,” which provide quantitative as well as qualitative recommendations on food intakes and lifestyle aspects, together constitute the FBDG for adults in Germany [54]. Regarding the food guide, few existing examples, including the Eatwell Guide [15] and those developed in Zambia and Rwanda following technical guidance by the Food and Agriculture Organization, have transferred recommended food group quantities from, for example, optimization models, as directly as presented in this study [16,17]. For the Wheel of Five in the Netherlands, the optimization results were first transferred into recommended amounts and only the results for adult women were used [11]. The transfer of food group quantities to servings, on the other hand, is one step often described within the development of FBDGs [56]. For the Dutch FBDG, diet optimization results were transferred in an iterative expert-based process [11]. For the Zambian FBDG, diet modelling results were combined with other findings on healthy diets to derive servings and recommendations [16]. In France, however, detailed background materials describe the derivation of healthy eating patterns via diet optimization, which provides the scientific basis for the FBDG, but information on the FBDG development process is not given [12]. Overall, a consistent system for the direct transfer of scientifically derived food quantities into FBDGs, as in our study, seems rare, particularly for the development of dietary messages.

Nevertheless, although consistency may strengthen the transparency of FBDGs, the specific implementation of our approach, which involved transferring optimization results into servings, may result in some simplifications (e.g. 12 g of egg/day corresponds to 1.4 eggs per week, which was rounded to 1 egg/week).

### Strengths and limitations

With the successful integration of diet-health relations, nutrient adequacy, and environmental and practical aspects into the diet optimization model, together with a consistent transfer process of the optimization results into the FBDG, the new methodology adheres to international frameworks for the derivation of FBDGs [3,6,57,58]. This approach forms the basis for accelerating and simplifying future update cycles for FBDGs. By using FoodEx2 as food codes, a Europe-wide applicable life cycle assessment dataset, and food intake data from the EFSA’s comprehensive database, our modelling framework is also applicable to other (European) countries, as demonstrated in Austria [18]. One strength of our approach is the simultaneous consideration of multiple dimensions. An alternative method to provide ideal food quantities for the derivation of FBDGs is via systematic reviews, as for example done for the Nordic Nutrition Recommendations. In that approach, the different dimensions (health and environment) were considered independently. Consequently, recommendations were provided separately, without an overall synthesis [55].

One limitation of our approach is the lack of science-based mitigation targets suitable for the context. To determine the magnitude of GHGE reduction, we used the mitigation goal of the Intergovernmental Panel on Climate Change [43]. It is rooted in science and represents a politically accepted global goal; however, it refers to all sectors globally. The most accurate way to define environmental goals may be to establish upper bound constraints linked to planetary boundaries. However, this approach requires careful consideration of the potential methodological differences between the applied European life cycle assessment data [37] and the methods used to derive planetary boundaries.

In general, the quality of our results is closely linked to the quality of the data used, in terms of currency, compliance with scientific standards, representativeness, and availability. The limitations were the age of the food intake data and, therefore, the inability to include plant-based alternatives to animal-source foods; the lack of diet–health relation data for all food groups; and the use of only two indicators (GHGE and land use) to assess environmental impact. Furthermore, certain aspects could not be operationalized in the optimization model because of a lack of data. For example, increased legume cultivation is associated with improved soil health [59]. Nevertheless, this information was incorporated into the recommendations, as we intentionally formulated them to recommend eating *at least* one portion of legumes per week. To maintain the best possible dataset, it should be updated regularly. For example, the EFSA is expected to continue to update the food classification FoodEx2 [60] and food intake databases [31].

## Conclusions

The FBDG 2024 for German adults are based on optimized diets that are nutrient-adequate, consider the habitual national diet, and substantially reduce the burden of diet-related disease and environmental impact. Consistent steps for transferring the optimization results into the DGE nutrition circle and messages increased the transparency of the derivation process. Our study showed that multi-objective diet optimization models have the potential to meet both current and future requirements for FBDGs. They will benefit from further research on e.g. clear mitigation goals for environmental impacts at the national level and on methods for the integration of future requirements, for example, regarding the food system. The diet optimization and transfer methods presented can be applied to specific target groups (e.g., children and adolescents, adults over 65 years, and vegetarians) and can be further developed with updated data and additional indicators (e.g., water use or contaminants).

## Data Availability

All data produced in the present study are available upon reasonable request to the authors.

## Acknowledgements

We thank Julia Haardt, Micha Limbeck, Margrit Richter, Christina Breidenassel, Manuela Michel, Maike Gutmann (German Nutrition Society), and Annika Blumenthal for their support in the development of the optimization model’s concept and the transfer process. We thank Anne Klatt, Hyewon Seo, Claudius Grehl, Ulrich Gromke (German Environment Agency), Hermann Lotze-Campen and Lisa Maria Pörtner (Potsdam Institute for Climate Impact Research) for their support in the determination of environmental indicators and goals. We thank Anita Frehner and Christian Schader (Research Institute of Organic Agriculture) for their support in the definition and interpretation of agronomic dependencies. We thank Romane Poinsot (MS-Nutrition) for her support of the modelling process.

ACS, HB, JC, UN, and BW designed research. KG, HH, AK, JL, SL, LS, and KV critically reviewed and discussed development of the optimization model and its outcomes. RG and FV provided the programming of the optimization model. ACS analyzed data. ACS, HB, and BW wrote paper. BW had primary responsibility for final content. All authors have read and approved the final manuscript.

## Data availability

Data described in the manuscript, code book, and analytic code will be made available upon request.

## Declaration of generative AI and AI-assisted technologies in the writing process

The authors declare that no generative AI or AI-assisted technologies were used in the writing of this manuscript.

## Abbreviations

DGE: German Nutrition Society (Deutsche Gesellschaft für Ernährung e. V.)
En%: Percentage of energy
EFSA: European Food Safety Authority
FBDGs: food-based dietary guidelines
GHGE: greenhouse gas emissions
WS: weighting scheme

