## Supplementary Methods for "Outcomes of a multi-objective diet optimization model as a basis for food-based dietary guidelines"

### Background

The update of the food-based dietary guidelines (FBDG) for Germany was commissioned by the scientific committee of by the German Nutrition Society (Deutsche Gesellschaft für Ernährung e.V., DGE) in 2016. The nominated FBDG working group was as followed: Prof. Dr. Bernhard Watzl (Chair), Prof. Ulrike Arens-Azevêdo, Prof. Dr. Heiner Boeing, Dr. Johanna Conrad, Prof. Dr. Kurt Gedrich, Dr. Maike Gutmann, Julia Haardt, Prof. Dr. Hans Hauner, Prof. Dr. Helmut Heseker, Prof. Dr. Anja Kroke, Prof. Dr. Jakob Linseisen, Prof. Dr. Stefan Lorkowski, Prof. Dr. Ute Nöthlings, Prof. Dr. Britta Renner, Anne Carolin Schäfer, Prof. Dr. Lukas Schwingshackl, and Dr. Kiran Virmani. From start (2016) until publication of the FBDG (2024), the working group had 22 meetings.

To gather information on current methods, the working group carried out literature searches on international approaches to derive FBDGs and organized a workshop on mathematical approaches jointly with the Federation of European Nutrition Societies on 23rd-24th September 2019 in Bonn [1].

### Concept development and method implementation

The working group developed a concept for the diet optimization model, which was transferred into code by MS Nutrition (Marseille, France; https://ms-nutrition.com/en/). After a process of testing and refinement, the DGE initiated a consultation workshop on 15th December 2022) on version 22.0.0 of the diet optimization model. The DGE invited scientific experts (nutrition, climate, environment) with the aim of having the new method evaluated by experts both within and outside the working group's field of expertise.

### Method refinement

Subsequent revisions (i.e. adaption of the environmental goal) led to version 23.0.0 of the optimization model, which was made available for commentary in a public consultation starting 13th March 2023 till 30th April 2023). A total of 1.094 comments were submitted by 66 individuals. All comments were summarized thematically and answered so that the implementation of the comments can be transparently tracked. The comments, answers, and declarations of conflicts of interest were published [2]. After further refinements based on the feedback from the consultation, the final version of the optimization model (version 24.0.0) was developed. The DGE invited stakeholders from nutrition counselling, community catering, and nutrition organizations to a workshop on 7th December 2023 to inform about and discuss the communication concept of the FBDGs. The final FBDG were published at the scientific congress of the DGE on 5th March 2024. Afterwards, the DGE organized an online information session on the new FBDG for the food industry and food retail sector on 7th March 2024.
